# Risk factors for pneumococcal carriage in adults living with HIV on antiretroviral therapy in the infant pneumococcal vaccine era in Malawi

**DOI:** 10.1101/2022.05.12.22274986

**Authors:** Deus Thindwa, Thandie S Mwalukomo, Jacquline Msefula, Kondwani C Jambo, Comfort Brown, Arox Kamng’ona, Charles Mwansambo, John Ojal, Stefan Flasche, Neil French, Robert S Heyderman, Todd D Swarthout

## Abstract

**Objective:** Adults living with HIV (ALWHIV) on antiretroviral therapy (ART) are at high risk of pneumococcal carriage and disease. To help evaluate carriage risk in African ALWHIV in the infant pneumococcal conjugate vaccination era, we assessed association between carriage and potential risk factors.

**Methods:** Nasopharyngeal swabs were collected from adults aged 18-40 years attending an ART clinic during rolling, cross-sectional surveys in Blantyre, Malawi between 2015-2019. We fitted generalised additive models to estimate the risk of sex, social economic status (SES), living with a child <5y, and ART duration on carriage.

**Results:** Of 2,067 adults, median age was 33y (range 28-37), 1,427 (69.0%) were females, 1,087 (61.4%) were in low-middle socio-economic-status (SES), 910 (44.0%) were living with a child <5y, and median ART duration was 3.0 years (range 0.004-17). We estimated 38.2% and 60.6% reductions in overall and vaccine-serotype carriage prevalence. Overall carriage was associated with low SES, living with a child <5y and shorter duration on ART. By contrast, vaccine-type carriage was associated with living without a child <5y and male sex.

**Conclusion:** Despite temporal reductions in overall and vaccine-serotype carriage, there is evidence of incomplete VT indirect protection. A targeted-vaccination campaign should be considered for ALWHIV, along with other public health measures to further reduce vaccine-serotype carriage and therefore disease.

## Introduction

*Streptococcus pneumoniae* (the pneumococcus) is a common coloniser of the human nasopharynx, particularly in young children and populations with human immunodeficiency virus (HIV) ^1^. Pneumococcal colonisation is a necessary prerequisite for transmission and the development of disease, including otitis media, sinusitis, pneumonia, meningitis, and bacteraemia ^2^. The pneumococcus is associated with a large burden of disease in adults living with HIV (ALWHIV) compared to adults without HIV ^3–5^. Adult HIV prevalence remains high (>10%) in many sub-Saharan African countries, with Malawi reporting a national prevalence of 10.6% ^6–8^. The use of antiretroviral therapy (ART) has substantially increased survival and reduced the incidence of invasive pneumococcal disease (IPD) ^9^. However, despite more than 85% of ALWHIV in Malawi receiving ART ^10–12^, ALWHIV remain at greater risk of IPD than adults without HIV ^3^.

Pneumococcal conjugate vaccines (PCVs) are widely used in infant schedules in low- and middle-income countries (LMICs), generally targeting the most commonly invasive serotypes in this age group ^1^. To date, in contrast to high-income settings, immunisation of vulnerable adults with pneumococcal vaccines has not been adopted in most LMICs ^13^. In November 2011, Malawi introduced the 13-valent PCV (PCV13) into the national Expanded Program on Immunisation (EPI) using a three-primary-dose schedule without booster (3+0; one dose at 6, 10 and 14 weeks of age). Despite nearly 10 years of >80% PCV13 three-dose coverage among age-eligible children, there is evidence of a sub-optimal reduction in both vaccine-serotype (VT) carriage prevalence and VT-IPD incidence in children and ALWHIV in Malawi ^14–16^. Similar evidence of residual VT carriage prevalence is also reported in the Gambia and Mozambique after 5 and 2 years of implementation, respectively, ^17^ despite both countries also reporting >80% PCV three-dose coverage under a 3+0 vaccine schedule ^18,19^. There is increasing evidence that the indirect protection (i.e. herd immunity) offered by an infant PCV against VT carriage, especially in ALWHIV, is sub-optimal ^14,18^.

The most effective strategy to reduce residual VT-IPD burden in ALWHIV depends on the factors shaping VT carriage and disease risk. Before introducing infant PCV in Malawi and South Africa, risk factors for IPD in ALWHIV included younger age, female sex, cotrimoxazole resistance, underlying medical conditions and living in a densely populated area ^3,4^. On the other hand, risk factors for carriage of any pneumococcal serotype included exposure to infants exposed to HIV ^20,21^, low socio-economic status (SES), high density living in housing with inadequate ventilation and with intense social contacts ^22–25^. Moreover, among Malawian ALWHIV, the overall (VT and non-VT [NVT]) carriage prevalence was found to be higher in those on ART than not on ART ^26,27^.

In the PCV era, however, there are important information gaps in our understanding of the relative importance of of key factors for pneumococcal carriage and disease among ALWHIV. These include duration on ART, PCV vaccination status among children in the household, and SES. Following our recent data from Blantyre, Malawi showing high residual VT carriage and its determinants in PCV13-vaccinated and unvaccinated children and in ALWHIV ^15,16^, we extend the analysis to identify age- and time-dependent risk factors for pneumococcal carriage in ALWHIV on ART using generalised additive modelling.

## Methods

### Study design

Blantyre is located in the southern region of Malawi spanning 2,025 km^2^, with an urban (population density 3,334/km^2^) and rural (253/km^2^) population of approximately 800,000 and 451,000 people, respectively. ^28^. As described elsewhere in more detail^15^, rolling, prospective cross-sectional pneumococcal nasopharyngeal (NP) carriage surveys were conducted between 29 June 2015 and 9 August 2019 in Blantyre to investigate the change over time of pneumococcal colonisation in ALWHIV on ART. The majority (98.6%) of sampled individuals were on a first line ART regimen containing either i) Zidovudine, Lamivudine and Efavirenz, ii) Tenofovir, Lamivudine and Efavirenz or iii) Tenofovir, Lamivudine and Nevirapine ^29^. Eight pneumococcal carriage surveys (each approximately 6 months in duration) were conducted from 3.6 to 7.9 years after infant PCV13 introduction into the EPI schedule. ALWHIV aged 18-40 years were recruited from the Queen Elizabeth Central Hospital (QECH) ART clinic in Blantyre using a systematic sampling approach. Exclusion from the study included being currently on treatment for tuberculosis, hospitalisation within two weeks of recruitment and previously enrolled in the survey.

### Nasopharyngeal sample collection and processing

An NP swab sample was collected from each participant and processed at the Malawi-Liverpool-Wellcome Programme laboratory, co-located to QECH, to ascertain the presence of pneumococci. Samples were collected and processed according to World Health Organisation guidelines ^30^.

Serotyping was done using latex agglutination, based on picking a single colony, to identify serotypes targeted by PCV13 (1, 3, 4, 5, 6A, 6B, 7F, 9V, 14, 18C, 19A, 19F, 23F). Non-typeable and NVT isolates were both classified as NVT. Pneumococcal carriage dynamics were further evaluated using DNA microarray techniques, a technique which, in the case of co-carriage of multiple pneumococcal serotypes, differentiates all individual serotypes and reports relative abundance of each serotype in carriage ^31,32^. Microarray was implemented only in surveys 1 through 4 and with samples having latex-confirmed pneumococcal carriage. Further details of sample processing has been reported earlier ^15,32^.

### Data collection and analysis

Participant data collected at recruitment included age, sex, cohabitation with a child <5y (Yes/No), social economic status (SES), duration of ART use, CD4+ T-cell count, current ART regimen and cotrimoxazole use. A multiple imputation random forest-based method, using MissForest R package ^34^, was conducted to impute 1 (0.0005%), 297 (14.4%), and 537 (26.0%) missing data points on cohabitation with a child <5y, SES, and duration of ART use, respectively. Though reported in the descriptive analysis, CD4+ cell count was excluded from model-based analyses because 46.0% of its data points were missing, a proportion above the acceptable standard threshold for imputation ^34^. Duration on ART was not used as a continuous variable because of data sparsity in age- or time-stratified analyses but was categorised as short (<3 years) or long (≥3 years) duration based on (1) a previous study in rural Malawi which showed strong evidence of high pneumococcal carriage during the first 2 years of ART use ^26^, and (2) the median value of ART duration in this study.

Individual fitted carriage prevalence estimates were categorised into 18-24y, 25-29y, 30-34y, and 35-40y age groups reflecting their distinct IPD incidence ^3^. Time was stratified into year of survey initiation (2015, 2016, 2017, 2018 and 2019). In Malawi, pneumococcal carriage prevalence is usually higher in the cold (May-August) and hot (September-November) seasons as compared to the rainy season (December-April) ^26^. Thus, seasonality in carriage was captured using an indicator variable with values ranging from January to December based on the month of NP sample collection.

### Generalised additive modelling framework

We used a generalised additive modelling (GAM) framework to fit to age- and time-specific trajectories of pneumococcal carriage, and allow flexibility in capturing nonlinear carriage dynamics. In brief, we used penalised B splines (P-splines) for the age and time spline smoothers to avoid knot selections which usually introduce under- and over-fitting biases when trading-off model fit to the data and the smoothness of the curve ^35^. A penalized log-likelihood maximization was used to fit a non-parametric binomial model with complementary log-log link function defined by log-hazard of carriage as a function of the risk factors and a spline in age and time. No time-series autocorrelation structure was included in the model fits because ALWHIV were independently sampled without replacement and with no evidence to suggest strong autocorrelation across time. Thus, the rolling, prospective cross-sectional carriage samples and model residuals were assumed to be serially independent.

### Age- and time-dependent carriage prevalence estimation

We modelled age- and time-dependent carriage trajectories separately for overall (VT+NVT) and VT carriage as outcome variables for a set of potential risk factors. Due to reported poor immunogenicity and effectiveness of PCV13 against serotype 3 ^36,37^, we also modelled VT carriage without serotype 3 (VT-st3) to explore changes in carriage prevalence. A model with age and time smoothers and potential risk factors including sex, seasonality, duration on ART, cohabitation with a child <5 years old, and SES were fitted to the carriage data to estimate the overall or VT carriage prevalence and risk factor-specific effects on carriage prevalence dynamics.

A ‘gam’ function in the ‘mgcv’ R package facilitated model fitting ^38^, based on a model formulated as *g*(*P*(*Y*_*i*_ = 1|*a*_*i*_, *t*_*i*_)) = *g*(*π*(*a*_*i*_, *t*_*i*_)) = *ŋ*(*a*_*i*_, *t*_*i*_) where *Y*_*i*_ is a binomial outcome on whether an individual *i* is carrying pneumococcus (1) or not (0); *g* is the complementary log-log link function; *π*(*a*_*i*_, *t*_*i*_) is the carriage prevalence estimate for individuals of age (*a*_*i*_) at time (*t*_*i*_); *ŋ*(*a*_*i*_, *t*_*i*_) is a nonparametric linear predictor as function of individual age and time, and a set of risk factors. The linear predictor on a predictor scale is further expanded using the equation *ŋ*(*a*_*i*_, *t*_*i*_) = *β*_0_ + ∑_*k*_ *β*_*k*_*G*_*i*_ + *te* (*a*_*i*_) + *te* (*t*_*i*_) where *β*_0_ is a model intercept, *G*_*i*_ refers to individual risk factor category *β*_*k*_, is the risk factor coefficient, *te* (*a*_*i*_) and *te* (*t*_*i*_) denote tensor product P-spline of predictor age (*a*_*i*_) and time (*t*_*i*_).

The relative difference in carriage prevalence was computed by subtracting the GAM carriage prevalence estimate for each age or time category from the reference category and then dividing the difference by the reference category and then multiplying by 100%. The 95% confidence interval (95%CI) of the relative difference was estimated using 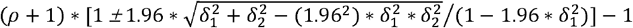, where *ρ* is the relative difference, *δ*_1_ and *δ*_2_ are the coefficient variations of the reference and comparator categories, respectively, and coefficient variation being standard deviation divided by the observed mean ^39^. GAMs with and without interactions between age group or time and each independent risk factor on the overall and VT carriage prevalence were fitted and compared using Akaike information criterion (AIC), and results of these tests are numerically presented in S1 Table. Given the model complexities, sensitivity analyses assesed factors that may affect carriage estimates which included the impact on carriage of individual age group or survey, serotyping method, carriage autocorrection, model formulation and spline type. Detailed sensitivity methods and results are presented in Supplementary Material (S1 Text and S2 Text). Analyses were conducted in R v4.1.1 ^40^, with statistical significance set at *p*<0.05, and the code is publicly shared via GitHub ^41^.

### Ethical approval

Ethical approval for this study was granted by the College of Medicine Research Ethics Committee, Kamuzu University of Health Sciences (P.02/15/1677), the Liverpool School of Tropical Medicine Research Ethics Committee (14.056) and the London School of hygiene and Tropical Medicine (26839). Individual written informed consent, including consent for publication, was obtained from each participant prior to study recruitment.

## Results

### Descriptive analysis

A total of 2,067 ALWHIV aged 18-40y were enrolled in the study between 29 June 2015 and 9 August 2019. Among adults with non-missing data, 1,427 (69.0%, n=2,067) were females, 413 (23.3%, n=1,770) and 674 (38.1%, n=1,770) were from low and middle SES households, respectively, 1,156 (56.0%, n=2,066) were not living with a child <5y, 1,772 (98.5%, n=1,799) were on one of Malawi’s first-line ART regimens, and 2,010 (97.2%, n=2,067) were using prophylactic cotrimoxazole at recruitment. The median age was 33y (IQR: 28-37, n=2,067), median CD4+ count was 252 cells/mm^3^ (IQR: 138-443, n=1,117), and median duration on ART at the time of study recruitment was 3.0 years, (range: 0-17, n=1,530) (Fig. 1).

**Figure 1.**
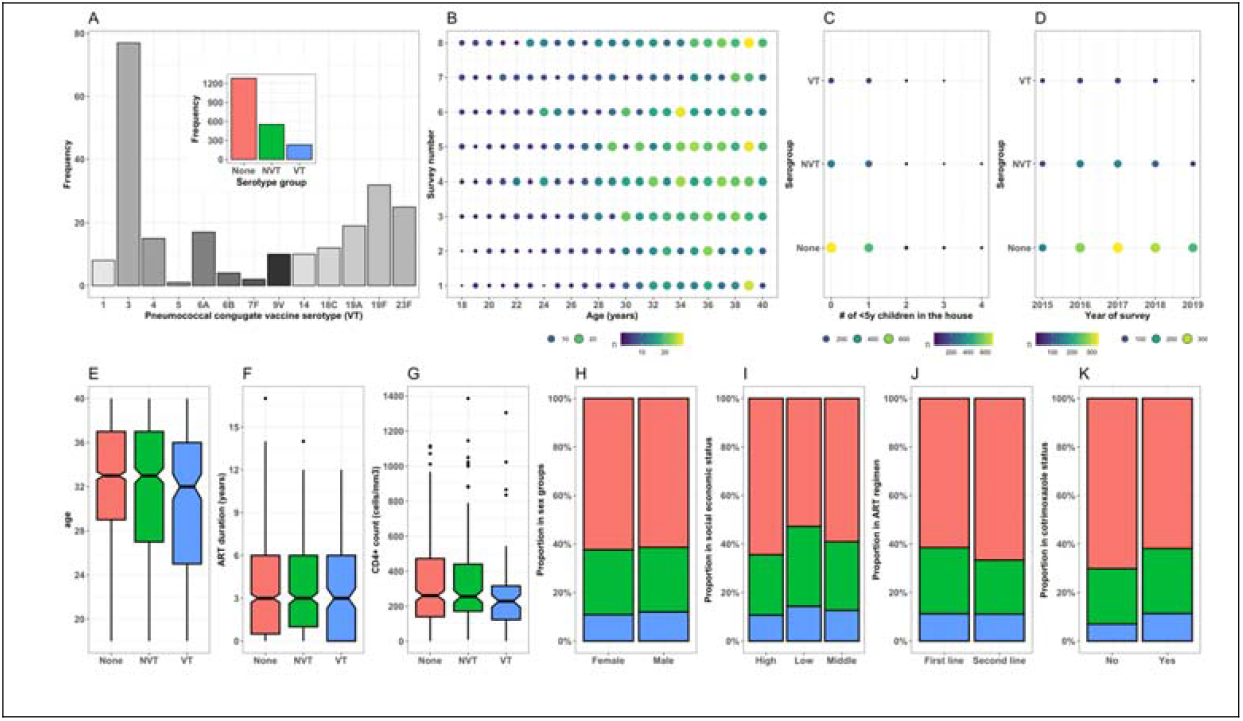
Demographics and clinical characteristics of participants using aggregated data across eight surveys. (A) Frequency of each VT in carriage; insert shows frequency of VT, NVT and no carriage. (B) Number of adults in each annual age per survey with circle size proportional to total sample size. The number of adults with VT, NVT, and no carriage living with (C) varying number of children <5 years or (D) across survey years. Notched box plots by serotype group representing participant distribution by (E) age, (F) duration on ART and (G) CD4+ count. Proportion of serotype group by (H) sex, (I) social economic status, (J) ART regimen and (K) Cotrimoxazole use.

Using survey-aggregated data, serotype 3 comprised 77 (33.2%, n=232) of all VT serotypes identified. Survey-aggregated data showed that carriage prevalence was 552 (26.7%, n=2,067) for overall (VT+NVT) and 232 (11.2%, n=2,067) for VT. It also showed that overall and VT carriage prevalence was 537 (37.7%, n=1,427) and 155 (10.9%, n=1,427) among females, 247 (38.6%, n=640) and 77 (12.0%, = 640) among males, 195 (47.2%, n=413) and 59 (14.3%, n=413) in low SES, 276 (40.9%, n=674) and 85 (12.6%, n=674) in middle SES and 243 (35.6%, n=683), 73 (10.7%, =683) in high SES households, 361 (39.7%, n=910) and 111 (12.2%, n=910) in adults living with a child <5y, 423 (36.6%, n=1,156) and 121 (10.5%, n=1,156) in adults living without a child <5y, 682 (38.5%, n=1,778) and 199 (11.2%, n=1,778) in adults on a first-line ART regimen, 9 (33.3%, n=27) and 3 (11.1%, n=27) in adults on second-line ART regimen, 767 (38.1%, n=2,010) and 228 (11.3%, n=2,010) in adults taking cotrimoxazole, 17 (29.8%, n=57) and 4 (7.0%, n=57) in adults not taking cotrimoxazole (Fig 1).

### Age- and time-dependent carriage prevalence estimates

Our GAM predicted a significant reduction in overall and VT carriage prevalence with increasing age and time. Among older age categories, overall carriage prevalence was lower than the reference younger adults aged 18-24y, with greatest reduction in adults aged 30-34y (−22.8%, 95%CI -34.1, - 10.4). Likewise, VT carriage prevalence was lower in older than younger adults, with highest reduction in adults aged 30-34y (−45.1%, 95%CI -61.8, -25.6). Across time, we estimated lower overall (−38.2%, 95%CI -51.7, -23.6) and VT (−60.6%, 95%CI -79.1, -39.2) carriage prevalence in 2019 compared to 2015. In a sub-analysis, serotype 3 made up 22.6-34.7% (across age groups) and 18.9-38.2% (across time) of VT carriage prevalence (Table 1; Fig. 2).

**Table 1.**
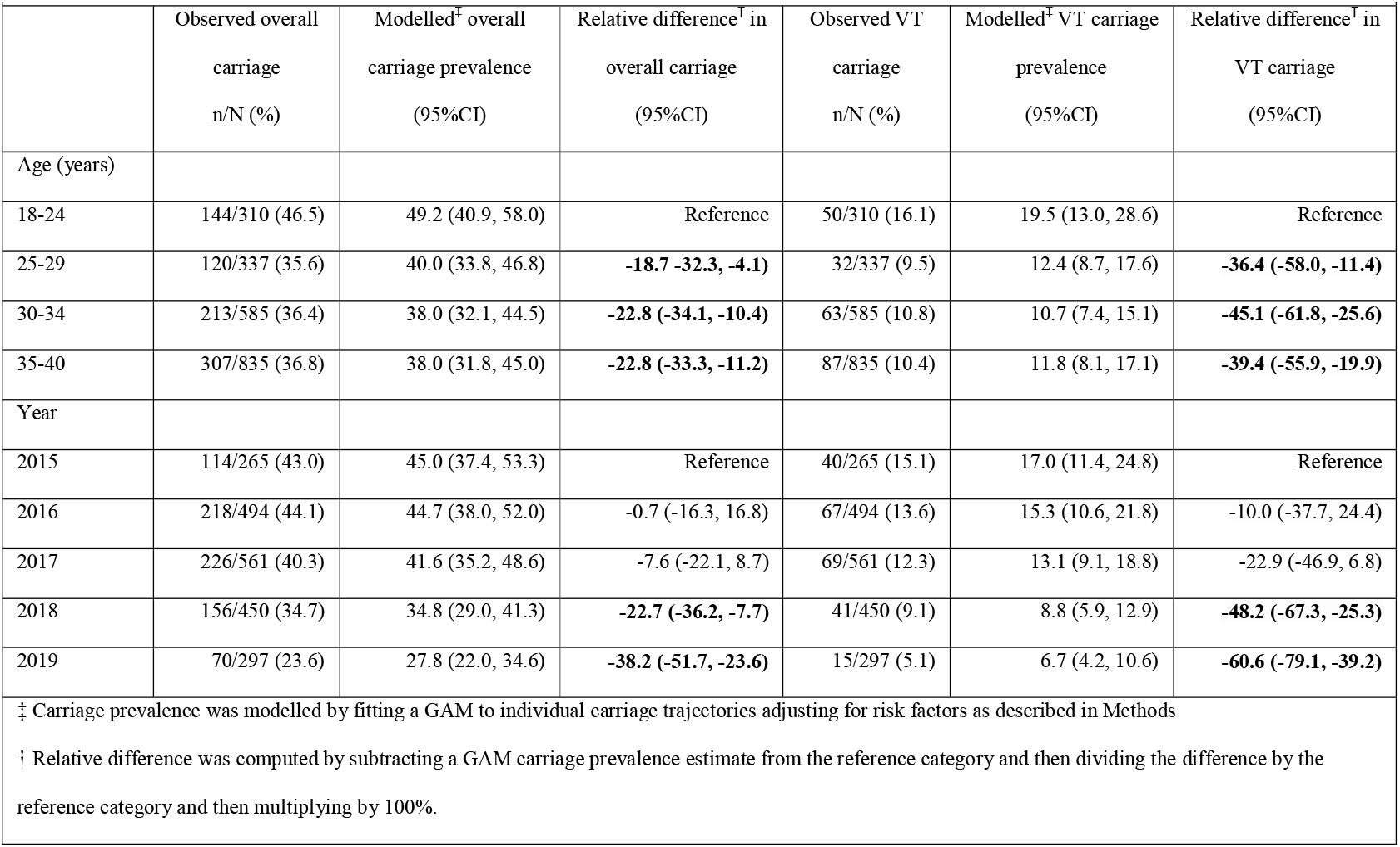
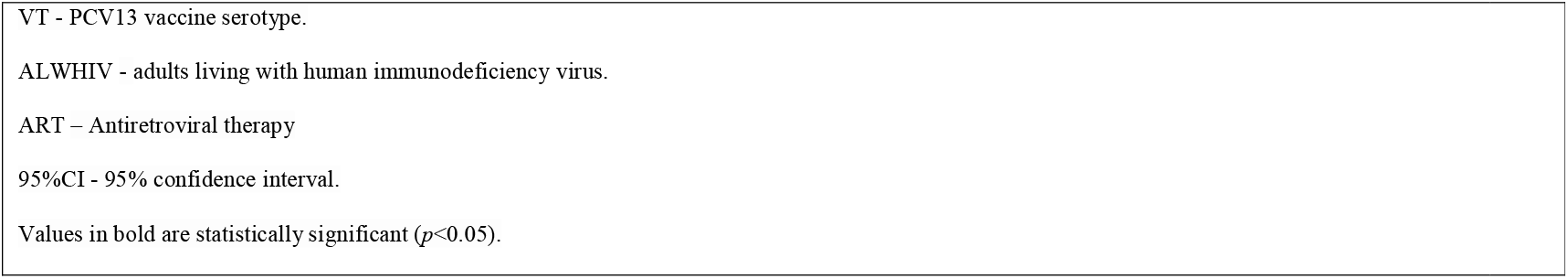
Age- and time-dependent overall and VT carriage prevalence, and relative difference in fitted carriage prevalence among ALWHIV on ART, 2015-2019 in Blantyre, Malawi.

**Figure 2.**
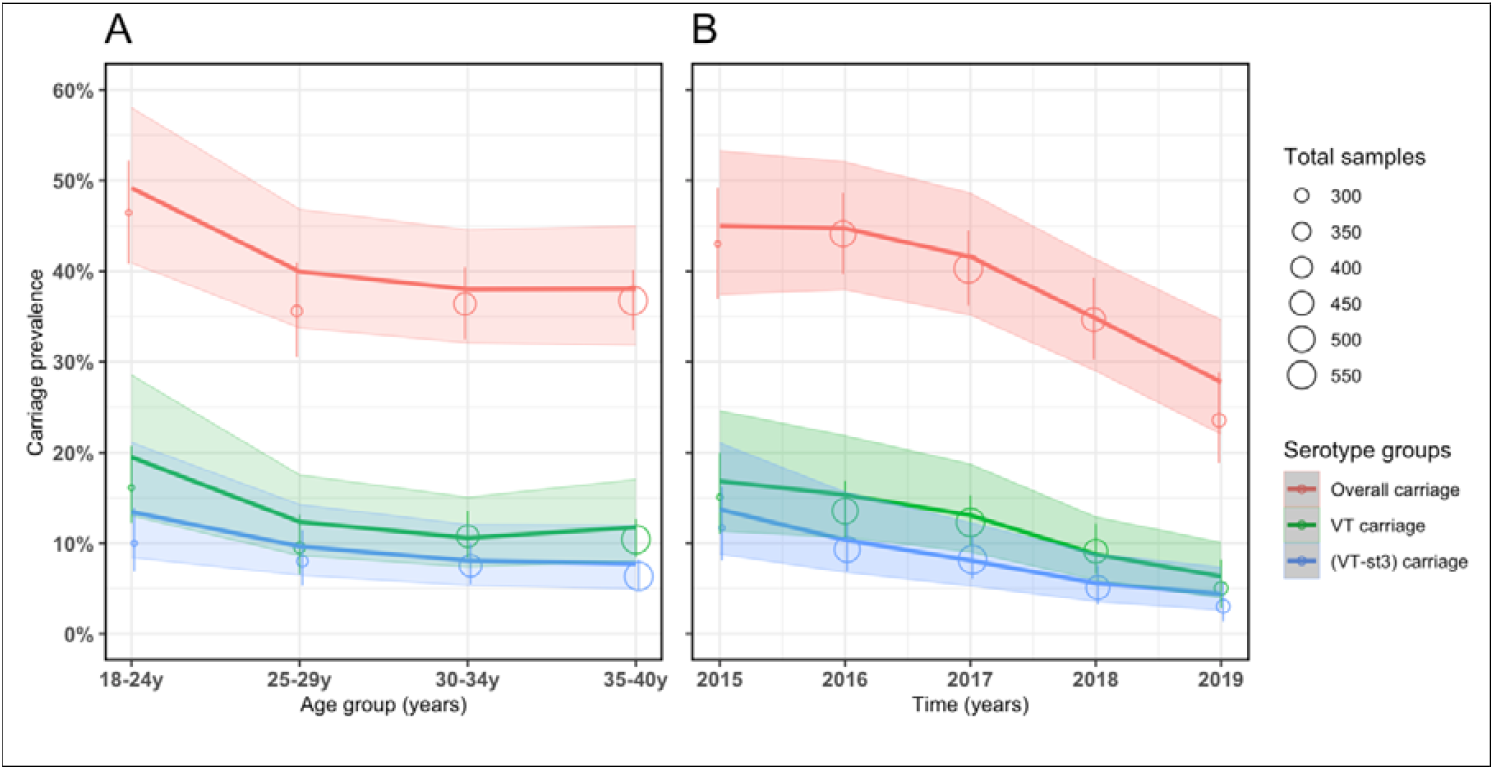
Observed and fitted pneumococcal carriage prevalence curves using data from rolling, prospective cross-sectional surveys in Blantyre, Malawi 2015-19. Number of carriage samples per age group between 18 to 40 years (y) and survey time from 2015 to 2019 represented by open circles radius proportion to total sample size with corresponding confidence intervals (vertical lines). P-spline GAM fitted lines and confidence intervals (ribbons) for the (A) age- and (B) time-dependent carriage prevalence stratified by overall carriage, vaccine serotypes (VT) carriage and VT carriage without serotype 3 (VT-st3).

### Factors associated with overall carriage prevalence

Overall carriage prevalence was only independently associated with SES, with adults in low SES having 22% higher overall carriage than those in high SES (21.9, 95%CI 1.6, 43.7). In a sub-analysis with age and time-stratification, our model predicted that being a younger (18-24y) adult in low SES or living with a child aged <5y was significantly associated with higher overall carriage prevalence. Significant associations with low SES and shorter ART duration were also seen with overall carriage. Overall carriage prevalence in younger adults was significantly higher by 42% for those in low vs high SES (41.8, 95%CI 12.5, 74.0) and 27% for those living with vs those living without a child <5y (27.2, 95%CI 0.4, 57.4). Temporally, overall carriage prevalence was persistently higher by 50% in 2018 (49.5, 95%CI 13.4, 89.8) and 84% in 2019 (83.6, 95%CI 20.5, 167.4) in adults in the low vs high SES, and higher by 35% in 2015 (35.4, 95%CI 0.9, 77.2) and 131% in 2019 (130.8, 95%CI 43.2, 255.4) in adults with shorter vs longer duration on ART (Table 2, Fig. 3, S1 Table).

**Table 2.** Risk factors for age- and time-dependent overall and VT with serotype 3 carriage prevalence, and the relative differences in the fitted carriage prevalence between the reference group and comparative groups among ALWHIV on ART, 2015-2019 in Blantyre, Malawi.

| Risk factors | Overall % relative difference<sup>†</sup><br>(95%CI) | % Relative difference<sup>†</sup> in age-dependent carriage prevalence (95%CI) |  |  |  | % Relative difference<sup>†</sup> in time-dependent carriage prevalence (95%CI) |  |  |  |  |
| --- | --- | --- | --- | --- | --- | --- | --- | --- | --- | --- |
|  |  | 18-24y | 25-29y | 30-34y | 35-40y | 2015 | 2016 | 2017 | 2018 | 2019 |
| Overall carriage |  |  |  |  |  |  |  |  |  |  |
| Female vs male | -4.8 (-20.9, 13.5) | -18.1 (-35.7, 1.7) | 16.3 (-15.9, 57.2) | 19.0 (-8.5, 52.9) | -10.3 (-25.5, 6.8) | -13.5 (-35.5, 12.7) | -6.1 (-23.9, 14.2) | 7.1 (-14.5, 32.5) | -12.9 (-33.8, 12.2) | 16.8 (-27.4, 79.3) |
| Low vs High SES | **21.9 (1.6, 43.7)** | **41.8 (12.5, 74.0)** | 2.1 (-26.0, 32.9) | 22.2 (-2.8, 49.2) | 19.9 (-1.8, 42.9) | 16.5 (-16.2, 52.2) | 19.3 (-5.4, 45.8) | 26.7 (-0.1, 55.2) | **49.5 (13.4, 89.8)** | **83.6 (20.5, 167.4)** |
| ART <3y vs ART ≥3y | 11.5 (-6.7, 31.5) | 1.9 (-20.8, 28.4) | 4.7 (-23.9, 40.6) | 4.4 (-15.7, 26.8) | 19.9 (-0.4, 41.8) | **35.4 (0.9, 77.2)** | 3.9 (-15.7, 25.3) | -0.7 (-19.4, 19.9) | 26.2 (-4.1, 61.4) | **130.8 (43.2, 255.4)** |
| With vs without child <5y | 9.7 (-0.8, 29.1) | **27.2 (0.4, 57.4)** | 2.6 (-22.9, 31.8) | 0.3 (-19.5, 22.1) | 8.9 (-9.2, 28.5) | 8.7 (-18.4, 39.5) | 1.1 (-17.9, 22.2) | 7.4 (-12.4, 29.4) | 13.8 (-14.3, 44.7) | -7.6 (-41.8, 31.8) |
| VT carriage |  |  |  |  |  |  |  |  |  |  |
| Female vs male | -18.6 (-64.9, 53.7) | -16.6 (-51.2, 28.6) | 26.8 (-42.8, 146.6) | -5.1 (-47.8, 59.1) | **-33.3 (-55.6, -5.8)** | -21.4 (-58.3, 31.3) | -18.0 (-48.9, 22.5) | -13.3 (-47.6, 33.1) | 1.1 (-54.4, 92.7) | 40.3 (-72.9, 315.6) |
| Low vs High SES | 6.0 (-61.0, 91.2) | -26.5 (-67.6, 19.6) | 12.5 (-46.6, 85.4) | 23.8 (-32.4, 90.8) | 35.8 (-14.5, 93.2) | 17.7 (-47.0, 94.6) | -3.2 (-48.7, 47.9) | 17.6 (-38.2, 80.9) | 37.3 (-39.2, 133.2) | 53.1 (-48.0, 229.1) |
| ART <3y vs ART ≥3y | 0.0 (-55.1, 77.4) | -14.5 (-49.6, 32.3) | -15.3 (-63.7, 65.3) | 33.7 (-19.3, 102.1) | -2.9 (-36.4, 35.0) | 32.7 (-26.2, 114.5) | -6.9 (-42.9, 36.2) | -21.9 (-51.5, 13.6) | 1.0 (-49.2, 67.6) | 54.4 (-53.7, 255.6) |
| With vs without child <5y | 33.1 (-40.9, 142.5) | **-40.2 (-64.7, -9.9)** | -6.6 (-52.3, 54.3) | 8.6 (-36.6, 67.6) | **-29.1 (-52.8, -0.5)** | 7.7 (-43.3, 73.9) | 26.9 (-21.2, 89.1) | 50.4 (-6.0, 125.0) | 4.5 (-55.1, 78.6) | **-84.8 (-108.2, -57.9)** |

<sup>†</sup> Relative difference was computed by subtracting a GAM carriage prevalence estimate of the reference category from the comparator category and then dividing the absolute difference by the reference category then multiplied by 100%.
ART: Antiretroviral therapy, ALWHIV: adults living with human immunodeficiency virus, CI: confidence intervals, y: year, SES: Social Economic Status score based a possession index which is calculated as a sum of positive responses for household ownership of each of the fifteen different functioning items such as watch, radio, bank account, iron (charcoal), sewing machine (electric), mobile phone, CD player, fan (electric), bednet, mattress, bed, bicycle, motorcycle, car, and television. Middle and high SES were combined and named as high SES.

**Figure 3.**
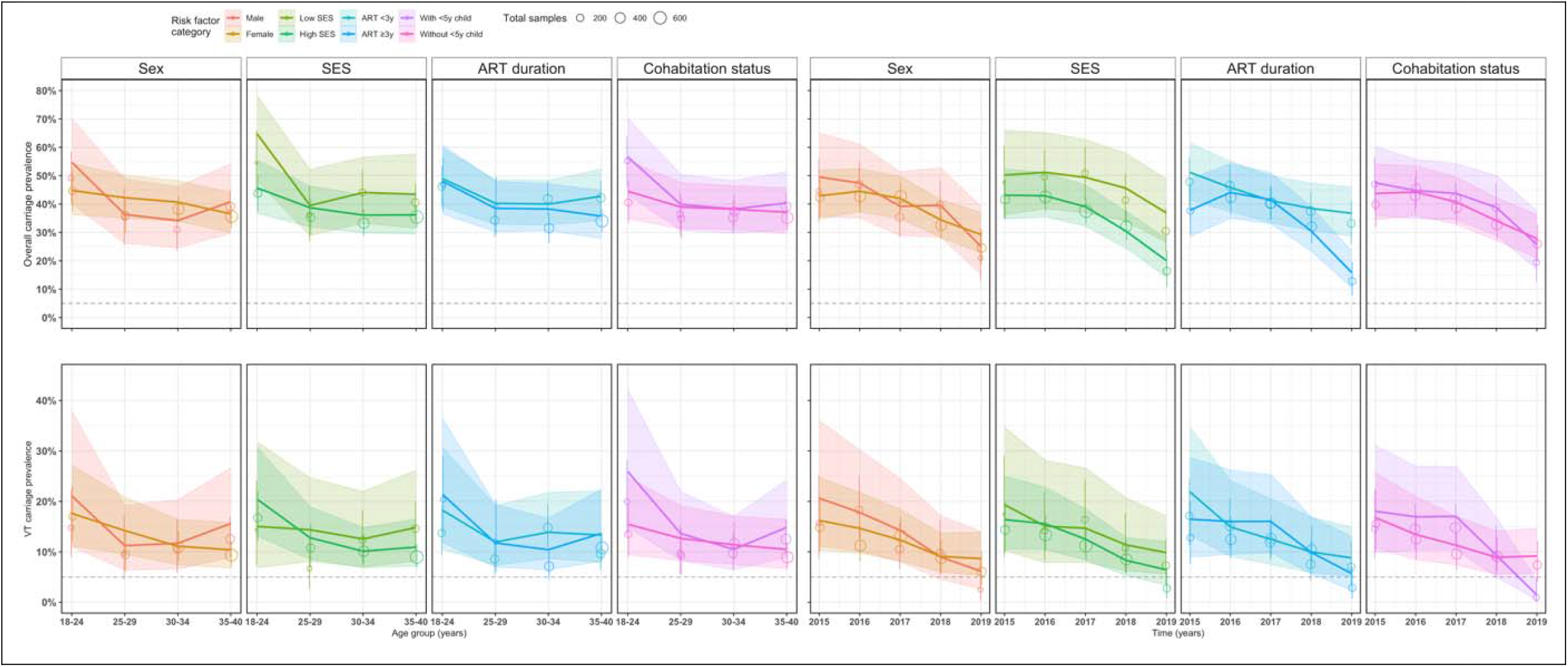
P-spline generalised additive model: Observed and fitted pneumococcal carriage prevalence curves for each potential risk factor category using data from rolling, prospective cross-sectional surveys in Blantyre, Malawi 2015-19. Nasopharyngeal samples across age groups from 18 to 40 years (y) and time represented by open circles. Circle radius is proportional to total sample size with corresponding confidence intervals (vertical lines). The coloured lines show P-spline GAM fitted model and confidence intervals for age- and time-dependent carriage prevalence for overall carriage (plots from first row) and vaccine serotypes (VT) carriage (plots from second row) stratified by risk factor categories.

### Factors associated with VT carriage prevalence

Sex, SES, ART duration and living with a child <5y were not significantly associated with VT carriage prevalence. However, with age- and time-stratification, our model of VT carriage outcome predicted that being a younger (18-24y) or older (35-40y) adult living without a child <5y or being older male significantly increased VT carriage prevalence. Temporally, living without a child <5y remained a significant predictor of higher carriage prevalence in 2019. Living without vs with a child <5y significantly increased VT carriage prevalence by 67% in younger adults 67.1 (95%CI 10.7, 140.5) and 41.0% in older adults (95%CI 1.5, 88.6). VT carriage prevalence was significantly higher in older males than females (50.0, 95%CI 7.2, 100.7; Table 2, Fig. 3, S1 Table).

## Discussion

We used generalised additive models to estimate age- and time-dependent overall (VT+NVT) and VT pneumococcal carriage prevalence and related risk factors in ALWHIV on ART. We analysed overall and VT carriage separately to take into account the effect of a high uptake infant PCV13 programme ^42^. Overall and VT carriage declined with increasing age group and time, with VT carriage having a faster decline (faster still if serotype 3 was excluded from VT). Our models predicted higher overall carriage prevalence in younger adults from low SES and living with a child <5y, as well as those with shorter duration on ART. Conversely, VT carriage prevalence was predominantly high in older males, and younger and older adults not living with a child <5y. These findings suggest that the decline in VT carriage prevalence across time in ALWHIV on ART is in part due to VT indirect protection from vaccinated younger children, although it is imperfect in males or adults not living with younger children, who may potentially have different routes of pneumococcal acquisition outside the household. Reduction in overall carriage suggests a combined effect of infant PCV13 vaccination and widespread use of antibiotics (i.e. cotrimoxazole) among adults in this setting.

Our analysis of risk factors for persistent carriage in ALWHIV on ART in the post PCV13 introduction era in urban Blantyre extends our previous observations that focussed mainly on high residual VT carriage and its determinants in PCV13-vaccinated and unvaccinated children ^15,16^. We now show substantially high overall and VT carriage prevalence in ALWHIV during the earlier (45% and 17%) than later (28% and 7%) years post infant-PCV13 introduction. VT carriage declined faster than overall carriage, suggesting cumulative vaccine-induced community-level indirect protection from infant PCV vaccination ^43–45^. The temporal reduction in VT carriage prevalence was even more marked when serotype 3 was excluded (and included as NVT), supporting accumulating evidence of the reduced effectiveness of PCV13 against serotype 3 ^36,37^.

Higher overall and VT carriage prevalences among younger than older adults reported in this study may suggest distinct high carriage acquisition risk in younger adults, partly supported by recent evidence of higher rates of skin-to-skin contacts between younger adults and with other age groups in urban Blantyre ^46^. The shorter median duration on ART amongst the younger adults as shown in S6 Fig may contribute to this residual pneumococcal carriage through incomplete immune reconstitution at both the systemic and mucosal level ^27,47,48^.

Low SES neighbourhoods in urban Blantyre predominantly comprise high-density informal settlements, relatively larger households and low rates of formal employment ^28^. Thus, substantial overall carriage prevalence in younger adults from low SES suggests that factors associated with low SES such as poorly ventilated and overcrowded houses with intense social contacts are reservoirs for pneumococcal carriage in the PCV13 era ^16,28,46,49^. On the contrary, non-differential VT carriage prevalence by household SES underlines an important role PCV vaccination plays to outweigh infection risks in poor settings. We uncover a phenomenon where adults living with children <5y, mostly PCV13 recipients given the high (>90%) infant PCV13 vaccination coverage ^15^, have substantially lower VT but higher overall carriage prevalence suggesting some non-vaccine serotype (NVT) replacement in adults within households, in line with evidence from rural Malawi and South Africa ^50,51^.

In this setting, VT carriage prevalence was higher in older male than female adults. VT carriage acquisition between mothers and their infants has been demonstrated previously in Malawi and South Africa prior to infant-PCV introduction ^20,21^. Thus, our finding aligns with recent evidence in the same setting showing strong intergenerational social mixing patterns between females and their potentially PCV13-vaccinated younger children likely through parental or guardian roles ^46^. This suggests that in the infant-PCV13 era, interruption of VT carriage transmission likely favors females than males.

Overall and VT carriage prevalence in ALWHIV on ART are heterogenous by age such that epidemiological models for carriage that incorporate ALWHIV should stratify for age for precise estimations. Our findings have policy implications in sub-Saharan African populations affected by HIV as persistent VT carriage in ALWHIV may imply continued risk of VT-IPD ^14^. The indirect impact on VT carriage of alternative infant-PCV13 vaccine strategies, including 2 primary doses with a booster dose or double booster doses (i.e. 2+1 or 2+1+1), currently being tested to improve the control of childhood disease, should also be further evaluated in ALWHIV ^18^. Indeed the 2+1 schedule, as implemented in South Africa, has generated indirect protection against IPD in unvaccinated older children and ALWHIV ^44,52^. However, simply improving control of carriage in young children to indirectly protect vunerable immunocompromised adults may be insufficient, particularly in the context of a high local force of infection and a rapid waning of vaccine-induced immunity ^16,53^. Furthermore, we provide evidence of heterogeneity in VT carriage prevalence with males or adults living without <5y child in their homes being at highest risk of VT carriage in the PCV era. Together, these data add weight to our viewpoint that as with many people living in high-income countries, targeted-pneumococcal vaccination should be considered in ALWHIV in LMICs.

We used a robust dataset with adequate samples to compute estimates for the overall, VT and risk factor-dependent carriage prevalence. Nonetheless, there were some limitations to our work, including limited data on risk factors such as viral load, use of tobacco, presence of other chronic co-morbidities, adherence to ART and history of antibiotics, which may independently influence carriage dynamics ^54^. In addition, latex agglutination method used in the main analysis for single serotype detection could underestimate our current prevalence estimates as compared to a more sensitive microarray method for multiple serotype detection as show in S2 Fig. Finally, the systematic recruitment of ALWHIV may be prone to bias if a cyclical pattern (unnoticeable here) is present in the important characteristics of the individuals as they attend the ART clinic ^55^.

In conclusion, despite temporal reductions in overall pneumococcal carriage, the risk of VT carriage and therefore subsequent pneumococcal disease remains high in ALWHIV. Efficient infant PCV schedules that enhance indirect protection together with targeted-vaccination for ALWHIV should be considered, along with other public health measures to further reduce VT carriage and disease. These measures should be supported by robust surveillance to assess effectiveness and identify early evidence of vaccine escape.

## Data Availability

The datasets generated during and/or analyzed during the current study are not publicly available, but are available from the corresponding author on reasonable request.

https://github.com/deusthindwa/Pneumo.carriage.adults.hiv.malawi

## Acknowledgements

We thank the individuals who participated in this study and the local authorities for their support. We are grateful to the study field teams (supported by Farouck Bonomali and Roseline Nyirenda). We are grateful to the hospitality of the QECH ART Clinic, led by Ken Malisita. Our thanks also extend to the MLW laboratory management team (led by Brigitte Denis) and the MLW data management team (led by the late Clemens Masesa whose contribution we wish to particularly acknowledge). DT, KCJ, JO, SF NF, RSH, and TDS are supported by the National Institute for Health and Care Research (NIHR) Global Health Research Unit on Mucosal Pathogens and RSH is a NIHR Senior Investigator. The views expressed in this publication are those of the authors and not necessarily those of the NIHR or the Department of Health and Social Care. The MLW Programme is supported by a Strategic Award from the Wellcome, UK

## Author contributions

Conceptualization; DT, SF, NF, TDS, RSH

Data curation; DT, TDS, TM

Formal analysis; DT, SF

Funding acquisition; NF, RSH, TDS

Investigation; DT, TM, AK, JM, CB, TDS

Methodology; DT, SF

Project administration; TM, AK, JM, CB, CM, NF, RSH, TDS

Resources; NF, RSH, TDS

Software; DT

Supervision; SF, NF, KCJ, TDS

Validation; DT, TM, KCJ, AK, JM, CB, CM, JO, SF, NF, RSH, TDS

Visualization; DT

Writing – original draft; DT

Writing - review & editing; DT, TM, KCJ, AK, JM, CB, CM, JO, SF, NF, RSH, TDS

All authors read and approved the final manuscript.

## Data availability

An R script that was used to analyse the datasets is available in the GitHub repository ^41^.

