## Supplementary file for "Risk factors for pneumococcal carriage in adults living with HIV on antiretroviral therapy in the infant pneumococcal vaccine era in Malawi"

**Title**

Corresponding author

### Supplementary Text 1

### Sensitivity analysis methods

In a sensitivity analysis, we first investigated the overall (VT+NVT) and VT carriage prevalence for separate age groups across time as well as for separate time points across age groups. Second, where microarray showed multiple serotype carriage, we estimated carriage prevalence comparing VT carriage based on latex serotyping (single serotype defined) to microarray serotyping that defined a sample as both VT and NVT if both were detected, irrespective of each serotype’s relative abundance. Third, to investigate potential differential smoothing parameters, we refitted the main P-spline model using three alternative types of splines including natural cubic spline, cubic regression spline and thin plate regression spline. Similarly, we evaluated the criteria for GAM selection by refitting the model (all risk factors were retained as before) with i) an age smoother only [$\beta_{0}+\sum_{k} \beta_{k}G_{i}+te\left( a_{i} \right)$], and ii) a time smoother only [$\beta_{0}+\sum_{k} \beta_{k}G_{i}+te\left( t_{i} \right)$], iii) both age and time smoothers [$\beta_{0}+\sum_{k} \beta_{k}G_{i}+te\left( a_{i} \right)+te\left( t_{i} \right)$], and iv) interaction between age and time smoothers [$\beta_{0}+\sum_{k} \beta_{k}G_{i}+te\left( a_{i} \right)+te\left( t_{i} \right)+te(a_{i},t_{i})$]. Finally, diagnostic checks for the P-spline model were conducted by checking residuals of the fitted models for autocorrelation of carriage samples across time.

### Supplementary Text 2

### Sensitivity analysis results

Given the model complexities and factors that may affect carriage estimates, sensitivity analyses were implemented to assess the impact on carriage of individual age group or survey, serotyping method, carriage autocorrection, model choice and spline type. Unlike VT carriage, a substantial 15.6-20.6% drop in overall carriage prevalence was seen in the last two surveys (2018-2019) compared to 2015-2017 surveys. Also, younger adults had a higher carriage prevalence (≥18.1% for overall and ≥28.6% for VT) than older adults across time (S1 Fig). Microarray was superior vs latex agglutination at detecting VT carriage, increasing detection by 6.3-11.4% across age groups and 6.4-10.6% across 2015-17 (S2 Fig). Residuals for GAMs fitted to overall and VT carriage for a set of risk factors were mostly random noise, with statistically non-significant autocorrelation in carriage time series (S3 Fig, S4 Fig). The choice of spline for GAMs had no significant influence on fitted carriage prevalence curves whereas the choice of model formulation had, with models that included both age and time splines having similar curves compared to those with only age or only time splines (S5 Fig).

| Supplementary Table 1. Test of interaction between age group or time and each independent potential risk factor on pneumococcal carriage prevalence using GAM models with and without interaction terms. | | | | | |
| --- | --- | --- | --- | --- | --- |
| Outcome (prevalence) | Interaction terms | DF | AIC for model without interaction | DF | AIC for model with interaction |
| Overall carriage | Age group*sex | 10.31861 | 2683.996 | 11.09804 | 2682.404* |
| Overall carriage | Age group*SES | 10.31861 | 2683.996 | 11.34193 | 2685.845 |
| Overall carriage | Age group*ART duration | 10.31861 | 2683.996 | 11.12247 | 2684.338 |
| Overall carriage | Age group*cohabitation^1^ | 10.31861 | 2683.996 | 11.26536 | 2682.175* |
| Overall carriage | Time*sex | 10.31861 | 2683.996 | 11.31885 | 2685.862 |
| Overall carriage | Time*SES | 10.31861 | 2683.996 | 11.39959 | 2684.419 |
| Overall carriage | Time*ART duration | 10.31861 | 2683.996 | 12.72311 | 2678.682* |
| Overall carriage | Time*cohabitation | 10.31861 | 2683.996 | 11.57051 | 2685.755 |
| VT carriage | Age group*sex | 9.609611 | 1431.703 | 10.62511 | 1432.768 |
| VT carriage | Age group*SES | 9.609611 | 1431.703 | 10.52064 | 1429.656* |
| VT carriage | Age group*ART duration | 9.609611 | 1431.703 | 13.42847 | 1427.124* |
| VT carriage | Age group*cohabitation | 9.609611 | 1431.703 | 10.63943 | 1431.334* |
| VT carriage | Time*sex | 9.609611 | 1431.703 | 10.70135 | 1431.697* |
| VT carriage | Time*SES | 9.609611 | 1431.703 | 10.77890 | 1432.731 |
| VT carriage | Time*ART duration | 9.609611 | 1431.703 | 10.84835 | 1431.816 |
| VT carriage | Time*cohabitation | 9.609611 | 1431.703 | 12.61655 | 1428.079* |
| VT - Vaccine serotype, DF - Degrees of freedom, AIC - Akaike Information Criterion, SES – socio-economic-status, ART – anti-retroviral therapy  * Model with interaction terms improves fit to the data  Cohabitation refers to participant living with one or more children <5 years old (Yes/No) | | | | | |

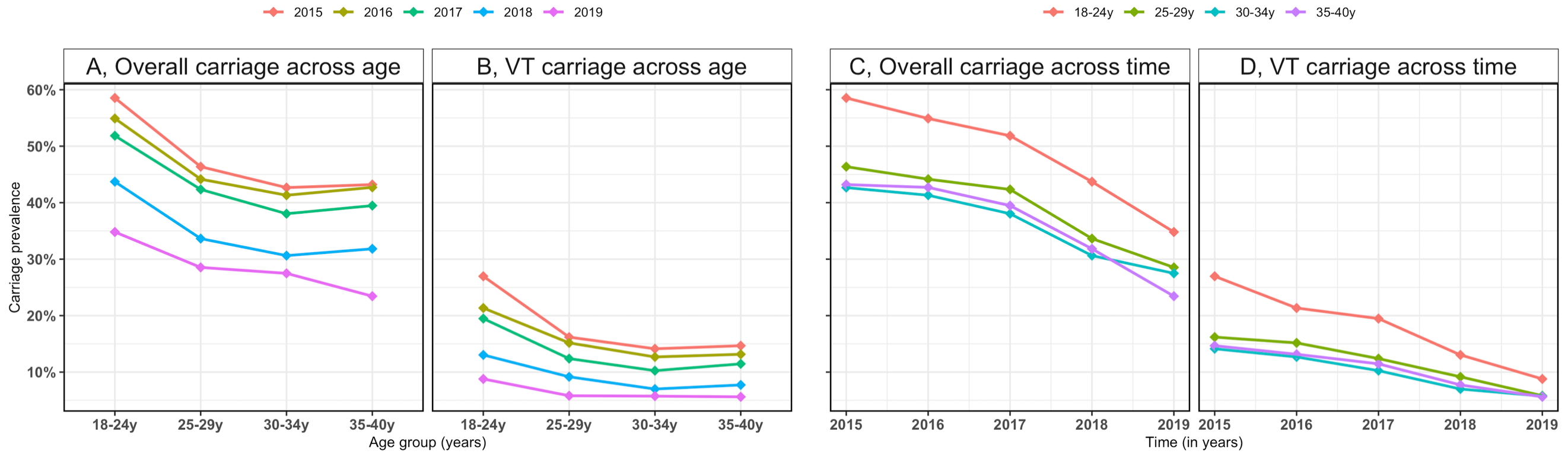

Supplementary Figure 1. Overall and vaccine serotype (VT) carriage prevalence for separate time points and age groups. (A) Overall carriage prevalence estimates for separate time points across age groups; (B) VT carriage prevalence estimates for separate time points across age groups; (C) Overall carriage prevalence estimates for separate age groups across time points; (D) VT carriage prevalence estimates for separate age groups across time points.

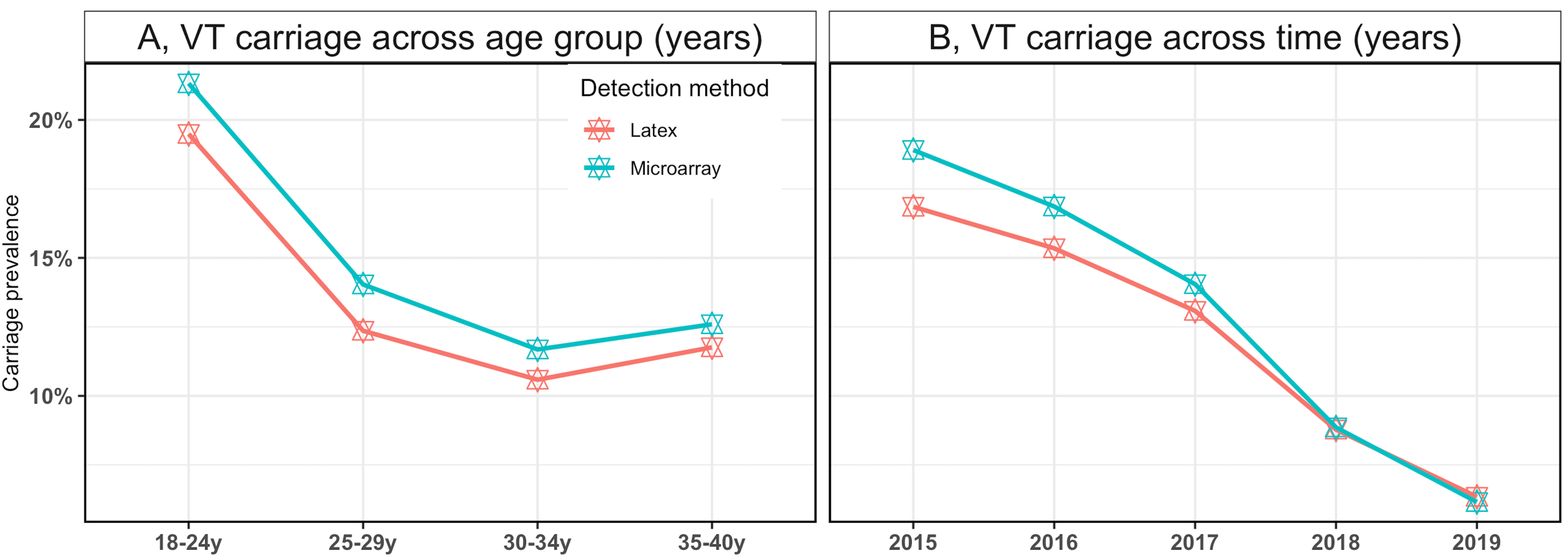

Supplementary Figure 2. Vaccine serotype (VT) carriage prevalence for two unique serotype detection methods across (A) age and (B) time. The latex agglutination method detected a single serotype, usually the one in highest abundance, whereas the microarray method detects all serotypes (VT and non-VT) carried and reports the relative abundance for each pneumococcal serotype in carriage. Assuming that each serotype contributes equally to carriage prevalence, detection by microarray has the advantage of detecting VT in low relative abundance and often not detected using latex agglutination, thus increasing VT carriage prevalence.

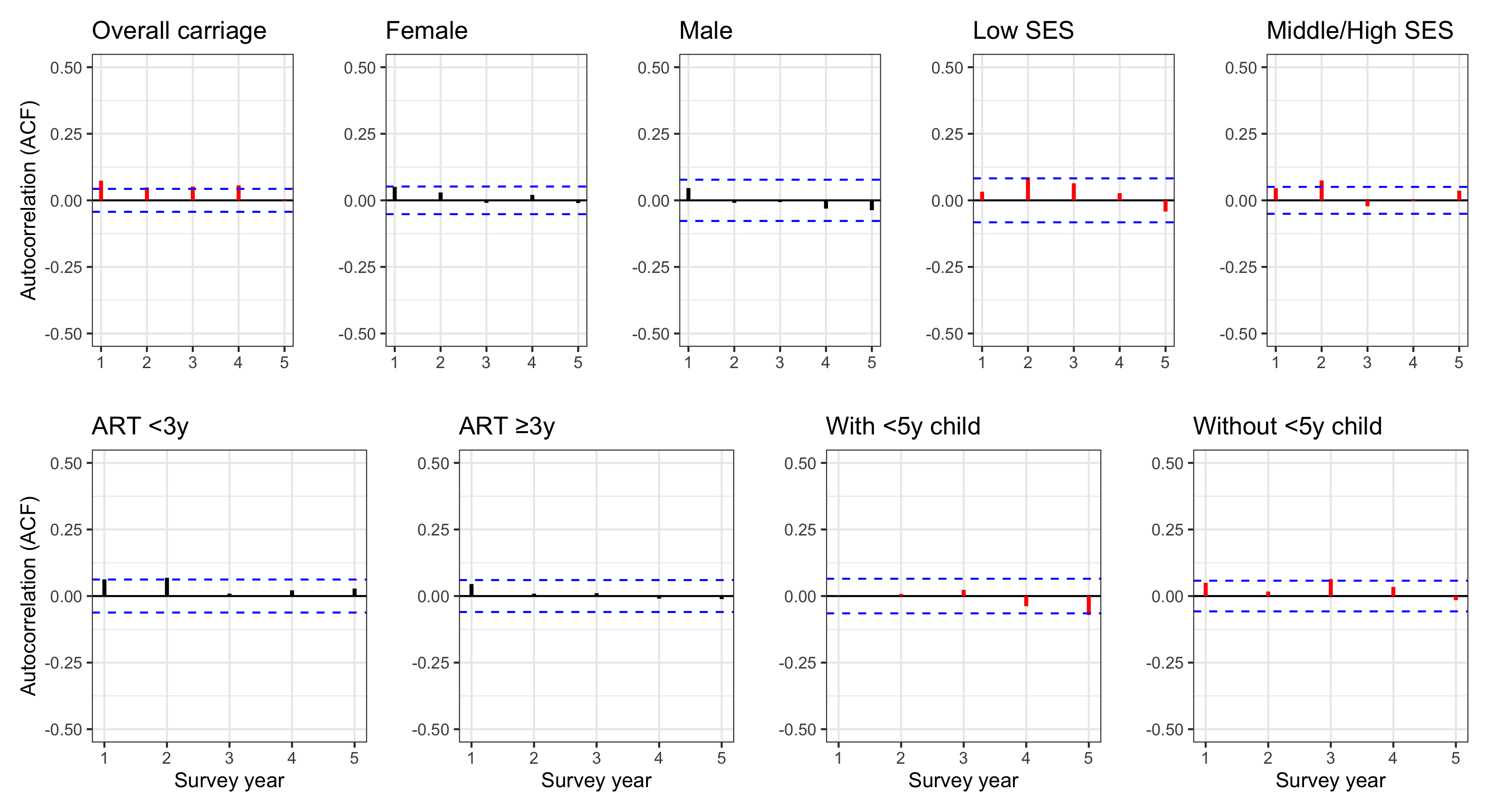

Supplementary Figure 3. Residual autocorrelation (ACF) plot of the main model of the overall carriage and related potential risk factors. No autocorrelation coefficients (values on the y axis) notably surpass the dotted blue region, implying that the autocorrelations across time are statistically zero at 95% confidence level, mostly representing random (white) noise.

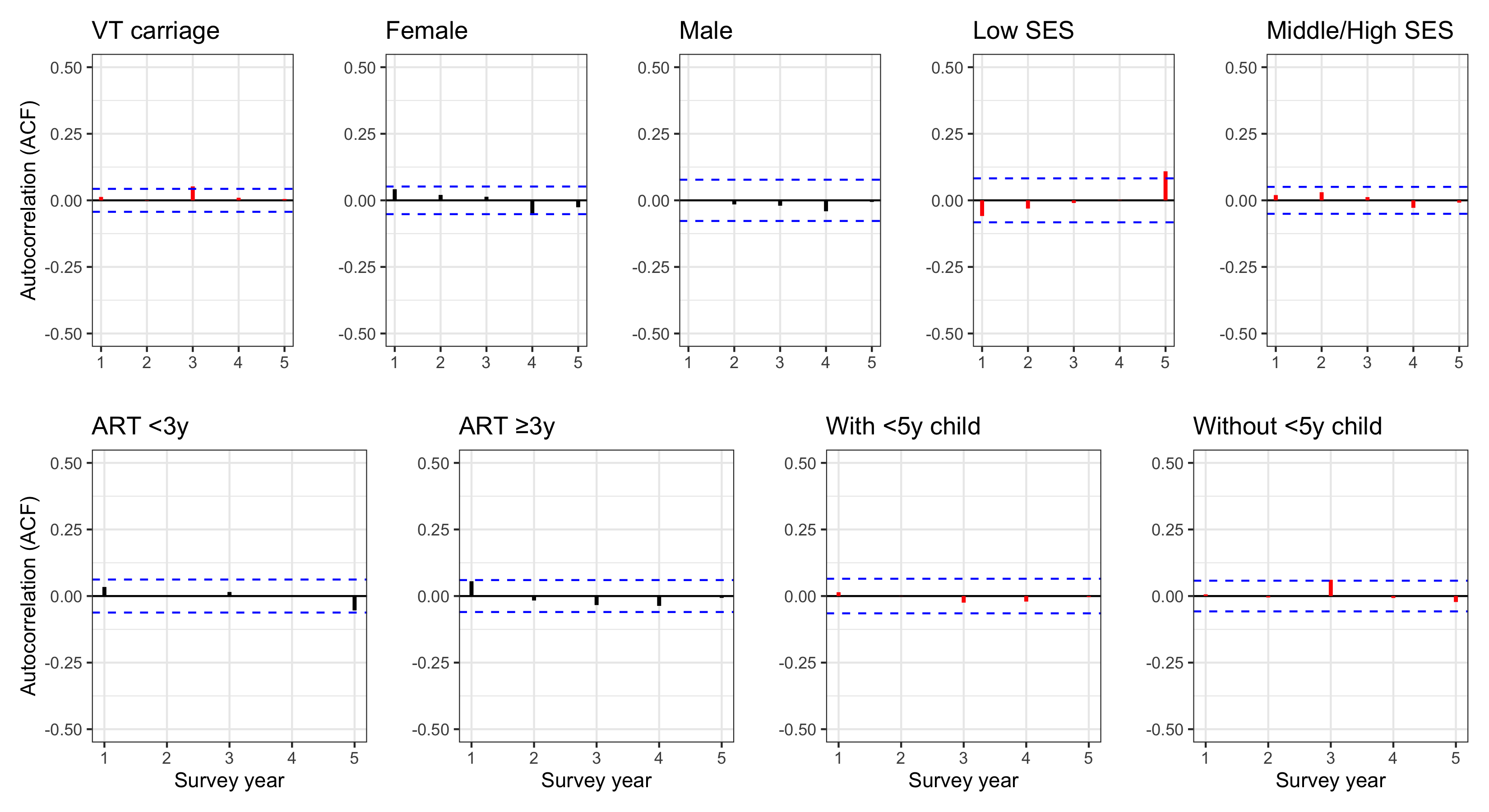

Supplementary Figure 4. Residual autocorrelation (ACF) plot of the main model of the vaccine serotype (VT) carriage and related potential risk factors. No autocorrelation coefficients (values on the y axis) notably surpass the dotted blue region, implying that the autocorrelations across time are statistically zero at 95% confidence level, mostly representing random (white) noise.

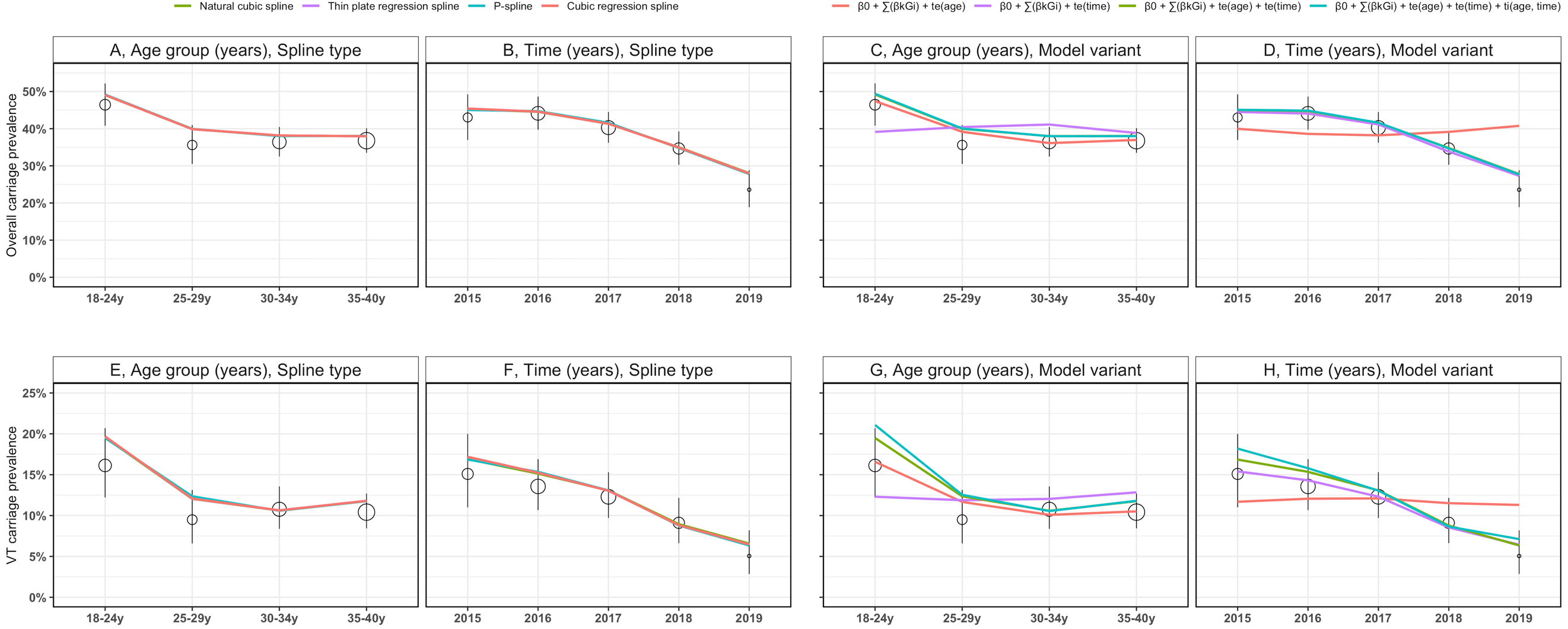

Supplementary Figure 5. Age- and time-dependent overall and vaccine serotype (VT) carriage prevalence by spline types and variants of model formulation. The left plots compare carriage prevalence curves for models that are fitted using natural splines, thin plate splines, P-splines and cubic regression splines to the overall carriage stratified by (A) age group, (B) time, and to the VT carriage stratified by (E) age group, (F) time. The right plots compare carriage prevalence curves for models that are fitted under different model formulations to the overall carriage stratified by (C) age group, (D) time, and to the VT carriage stratified by (G) age group, (H) time where $\beta_{0}$ is a model intercept, $G_{i}$ refers to individual risk factor category, $\beta_{k}$ is the risk factor coefficient, $te\left( age \right)$ and $te\left( time \right)$ denote tensor product P-spline of predictor age and time, respectively.

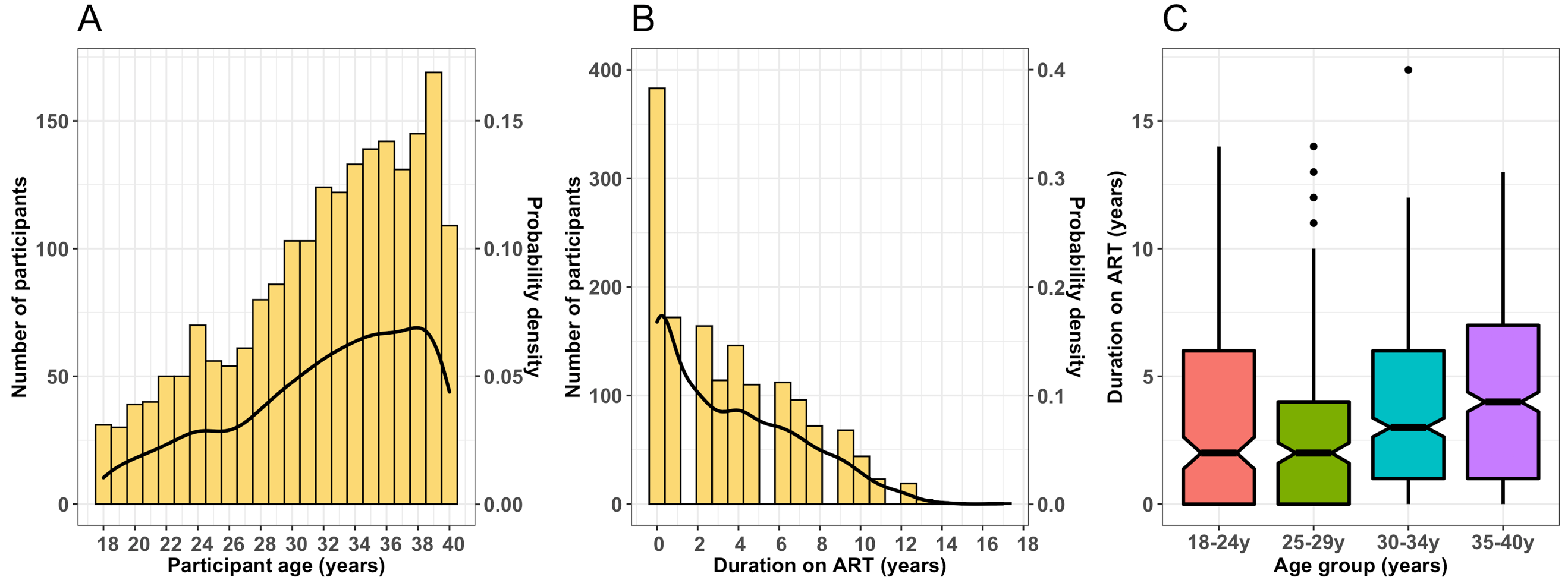

Supplementary Figure 6. Distributions of participant age and their duration on antiretroviral therapy (ART) in years. Histogram and density plots showing frequency (bars) and probability density (black line) of study participants by (A) age and (B) duration on ART, and (C) the stratification of duration on ART by age groups. The notched plots in (C) show median, minimum, maximum, interquartile range and 95% confidence intervals for the median of the duration of ART.
